# Improving Glioblastoma Classification Using Quantitative Transport Mapping with a Synthetic Data Trained Deep Neural Network

**DOI:** 10.64898/2026.03.31.26349864

**Authors:** Dominick Romano, Alexandra G Roberts, Benjamin Weppner, Qihao Zhang, Maneesh John, Renjiu Hu, Mert Şişman, Ilhami Kovanlikaya, Gloria C Chiang, Pascal Spincemaille, Yi Wang

## Abstract

**Purpose:** To develop a deep neural network-based, AIF-free, perfusion estimation method (QTMnet) for improved performance on glioma classification.

**Methods:** A globally defined arterial input function (AIF) is needed to recover perfusion parameters in the two-compartment exchange model (2CXM). We have developed Quantitative Transport Mapping (QTM) to create an AIF-independent estimation method. QTM estimation can be formulated using deep neural networks trained on synthetic DCE-MRI data (QTMnet). Here, we provide a fluid mechanics-based DCE-MRI simulation with exchange between the capillaries and extravascular extracellular space. We implemented tumor ROI generation to morphologically characterize tissue perfusion. We compared our QTMnet implementation with 2CXM on 30 glioma human subjects, 15 of which had low-grade gliomas, and 15 with high-grade glioblastomas.

**Results:** QTMnet outperforms (best AUC: 0.973) traditional 2CXM (best AUC: 0.911) in a glioma grading task.

**Conclusion:** The AIF-independent QTMnet estimation provides a quantitative delineation between low-grade and high-grade gliomas.

## Introduction

Glioblastoma is the most malignant form of primary brain tumors (1,2). As such, glioblastoma detection and grading is of paramount importance to disease prognosis and treatment planning (3,4). Dynamic Contrast-Enhanced MRI (DCE-MRI) has become an important tool in assessing glioblastoma (5-8). Although signal parameters can be used to grade glioblastoma (7), the findings are typically graded with tracer-kinetics modelling due to the connections to tissue physiology, as well as improved robustness and performance (9,10). Such techniques include the Extended Tofts Model (5,6) and its two compartment exchange (2CXM) generalization (11,12). These techniques rely on the selection of a global arterial input function (AIF), which leads to delay and dispersion artifacts and an issue in selection (13,14). The AIF dependence is shown to introduce variability in parameter estimation and classification performance (14-17).

Quantitative Transport Mapping (QTM) and other convection-diffusion based methods were proposed to overcome the AIF selection issue (13,18-20) by modelling the spatiotemporal variation of the contrast agent in relation to fluid velocity and diffusivity fields (13,18,19). QTM velocity estimations have outperformed AIF-based methods in several clinical tasks (18,21-24), however flow, exchange, and volume parameters are also desirable quantities for characterizing disease (11). In the QTM framework, along with other fluid-mechanical approaches, flow estimation requires knowledge of the underlying vasculature (25); however, this information is not available in DCE-MRI. To this end, we developed a framework that synthesizes vasculature and concentration data from randomly generated perfusion maps (15,25). The synthetic data is used to train a deep neural network (QTMnet) that learns the relationship between the concentration profile and perfusion parameters (15,25,26).

Previous work has emphasized the importance of matching the synthetic data domain to the in-vivo image domain (15,27), in which the disagreement between the synthetic training data and in-vivo data is described as domain shift (28,29). Several works address the domain shift problem by using large and highly varied training datasets (29-31). However, the transport forward problem is costly (18,25), and current attempts at generating enough training data for sufficient randomization are spatially and temporally untenable. Previous implementations have used uniform perfusion ROIs for training (15,25). We propose that including synthetic tumors, along with viable tumor perfusion states, will improve QTMnet performance. To this end, we have extended the pipeline to include various tumor and tissue perfusion ROIs within the simulation and consider exchange effects between the capillary and extravascular extracellular space (EES). We evaluated the performance of QTMnet and 2CXM parameter estimations on 30 glioma subjects.

## Methods

### Synthetic Tumor Perfusion Pipeline

We provide an overview of the previously described (15,23,25,32) tissue simulation pipeline. The process can be broken down into three steps: perfusion parameter label generation, vascular tree construction, and tracer transport simulation. The parameter maps are defined for a 32 × 32 × 32 *mm*^3^ volume. The simulation is implemented over a voxel grid with position vector 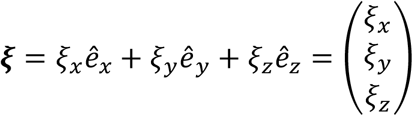. As in the previous work, we used a simulation resolution *dx* = *dy* = *dz* = 1*mm*. The corresponding voxel element is then *dV* = *dx* ⋅ *dy* ⋅ *dz* = 1*mm*^3^. In the current and previous works, a mixed Gaussian distribution was used to assign spatial variations within the volume of interest (15,25). The flow label *F* is used to construct the arterial and venous trees via Constrained Constructive Optimization (CCO) (33). A capillary distribution is generated in each voxel. Each segment has a flow and volume that will add up to the specified total voxel flow and volumes. Then, the transport simulation is run.

First, a boundary condition is modeled by a bolus function. This bolus function has its parameters drawn from a distribution and the curve is simulated. It is propagated down the tree using a parabolic velocity profile at each segment to calculate the transport of tracer along the length of the segment. Next, once the boundary bolus is propagated to the capillary network, we numerically solve the governing transport ODE for each capillary element. The capillary elements are combined using conservation of mass for each voxel. Finally, the capillary tracer mass is converted back to concentration for the venous simulation. In the venous tree, the tracer bolus is allowed to drain up to the venous root. The transport is also facilitated by parabolic flow in the segments (15).

The synthetic tumor perfusion pipeline builds onto the original QTMnet in the following ways: varying the simulation ROI, directly modeling tumor perfusion states, and extending the capillary simulation to include tracer exchange effects. Previous works (15,25) utilized synthetic cubes with a uniform ROI (**Figure 1**, right column). However, the CCO algorithm can be applied to a wide range of convex bounding geometries (33,34). This allows for the inclusion of tumor and tissue ROIs within the cubic simulation FOV.

**Figure 1.**
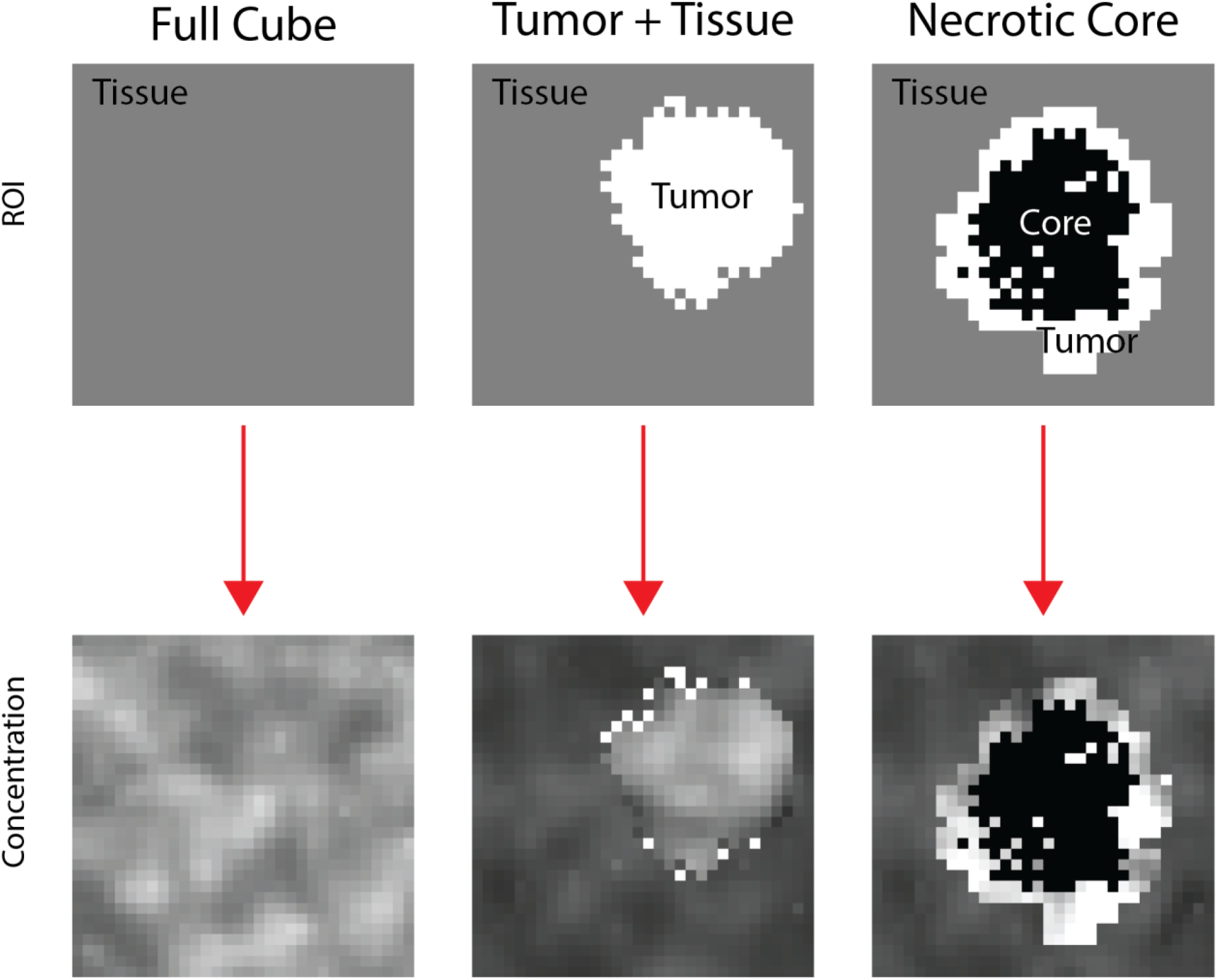
Overview of ROI definitions for tumor generation. (Left) A uniform ROI based on previous work. (Middle) A tumor ROI is generated with a region growing algorithm and additional dilation. The normal appearing tissue is the complement of the tumor ROI. (Right) A simulated tumor with necrotic core. To generate the core, an initial tumor ROI is made. Then, the initialized ROI is dilated. The core is defined as the initial tumor ROI, while the enhancing tumor ROI is the complement of the dilated ROI and the initial ROI. Finally, the normal appearing tissue ROI is defined as the complement of the dilated ROI.

To explicitly model the tumor, we split this cubic ROI into two complementing ROIs: the normal appearing tissue ROI and the tumor ROI. To generate the tumor ROI, we implemented a region growing algorithm. The algorithm has a fixed growth probability of *p*_*grow*_ = 0.5 and minimum ROI size of *N*_*min*_ = 163 corresponding to 0.005 ⋅ *N*_*FOV*_. Note that *N*_*FOV*_ corresponds to the total number of voxels in the simulation field of view, *N*_*FOV*_ = 32^3^. The growth probability was chosen due to the types of morphologies generated. *N*_*min*_ was implemented to ensure a viable ROI for training. For more details, see supplemental materials.

We allowed the number of seeds and maximum points *N*_*max*_ to vary to represent multiple, diverse tumors within a cube. The number of seeds can correspond to increasing the variability of tumor morphology and including the potential for multiple tumors to exist within the simulation FOV. We allowed the following number of seeds in this simulation: 1 (selection probability = 95%), 2 (selection probability = 3%), 3 (selection probability = 1%), 4 (selection probability = 1%). In accordance with the number of seeds, we allowed the maximum allowable tumor ROI points to vary between *N*_*max*_ = [0.05 ⋅ *N*_*FOV*_, 0.35 ⋅ *N*_*FOV*_]. With a higher seed number, a larger portion of *N*_*FOV*_ is allowed to be allocated to the tumor ROI.

The region growing algorithm acts as an initialization for the desired tumor ROI. In all cases, the region grown tumor ROI is dilated. In the solid tumor case (*p*_*solid*_ = 0.4), the region grown ROI is dilated by 2 (*p* = 0.3) or 3 (*p* = 0.7). The solid tumor ROI is then the resulting ROI after dilation, which can be called *M*_*tumor*_. Then the normal appearing tissue ROI, called *M*_*tissue*_, is the complement of the solid tumor ROI (**Figure 1**, middle column). In the remaining situations, we devised a simple method to implement a necrotic core (*p*_*core*_ = 0.6). To create the core mask, a region grown ROI is dilated by one voxel. Then, the core mask is dilated by an additional 3 to 6 voxels to make a dilated ROI. The tumor ROI (*M*_*tumor*_) is the complement of the dilated ROI and core ROI. To be clear, the core ROI is excluded from CCO and concentration simulation. Then, the tissue ROI (*M*_*tissue*_) is the complement of the dilated tumor mask (**Figure 1**, right column). The resulting ROIs (*M*_*tissue*_ & *M*_*tumor*_) are then individually fed into the CCO algorithm to obtain the arterial and venous trees. After CCO, the concentration simulation is played for the tumor and tissue ROIs.

The single compartment pipeline in (15) is extended to two compartments in the following way. We use a mixed Gaussian distribution to assign flow *F*, permeability surface product *PS*, capillary volume *V*_*p*_ and extravascular extracellular space volume *V*_*e*_. Literature values are used to guide the scale of perfusion parameters for normal appearing tissue (35,36) and tumor tissue (37,38). For the capillary simulation in normal appearing tissue, *N*_*cap*_ capillaries are drawn from a uniform distribution such that *N*_*cap*_ ∈ [50,120] for each simulation FOV. In the tumor tissue, the uniform distribution is defined such that *N*_*cap*_ ∈ [50,305]. *F, PS* & *V*_*p*_ are then assigned to a distribution over the *N*_*cap*_ capillary elements, which will be denoted *F*(***𝝃***, *ζ*_*c*_), *PS*(***𝝃***, *ζ*_*c*_) & *V*_*p*_(***𝝃***, *ζ*_*c*_) We define ***𝝃*** as the voxel coordinate and *ζ*_*c*_ ∈ [1,2, …, *N*_*cap*_] as the capillary index (13,15,25). The samples of the distribution are scaled to ensure that the assigned voxel values are recovered, meaning

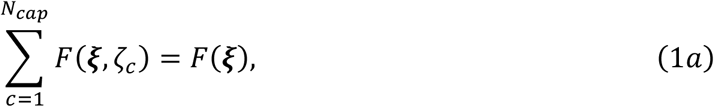

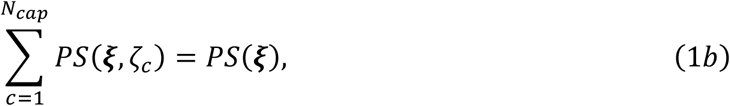

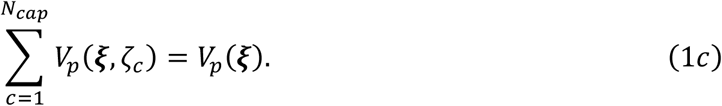

The boundary condition is a critical component to successful implementation. For brain DCE, we used the Parker Model as our boundary condition at voxel coordinate ***𝝃***_*root*_ (39):

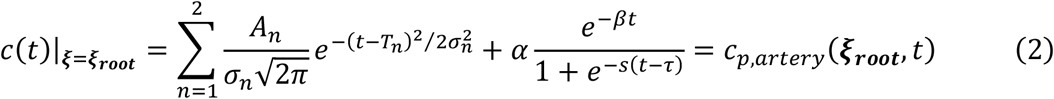

Where we draw the Parker model parameters *ρ*_*i*_ ∈ {*A*_*n*_, *σ*_*n*_, *T*_*n*_, *α, β, s, τ*} from a normal distribution such that 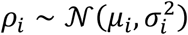, where *μ*_*i*_ is the parameter mean and *σ*_*i*_ is the parameter standard deviation. For a more in-depth explanation of *ρ*_*i*_ and parameter values, see corresponding supplemental section and table **T1**. These parameters were based on Parker’s reported values and our dataset considerations. The boundary condition is propagated through the arterial network to obtain the arterial concentration *c*_*p*,*artery*_(***𝝃***, *t*), and to deliver the tracer to the capillary inlet as done in previous work (15). At each voxel coordinate ***𝝃*** = ***𝝃***_***term***_, we call this the arterial terminal concentration, or *c*_*p*,*artery*_(***𝝃***_*term*_, *t*). The capillary inlet concentration is then set to the arterial terminal concentration:

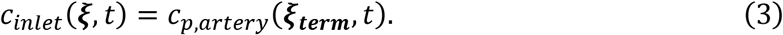

We now have what we need to simulate capillary transport.

In the capillary network, we implement a two-compartment model of transport and exchange. In the capillary space, we have concentration *c*_*p*_(***𝝃***, *ζ*_*c*_, *t*) (units: *mmol*⁄*mm*^3^). The extravascular extracellular space has tracer *mass C*_*e*_(***𝝃***, *ζ*_*c*_, *t*) (units: *mmol*). In our simulation, *F* is in units *mm*^3^⁄*s, PS* is in units *s*^−1^, and *V*_*p*_, *V*_*e*_ are in units *mm*^3^. With initial conditions

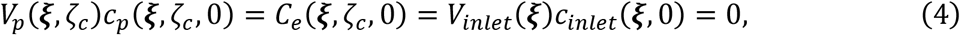

We use Euler’s method to numerically solve for the following mass balance system of differential equations

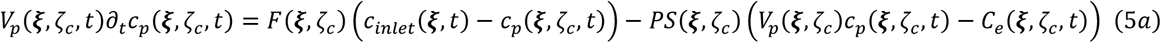

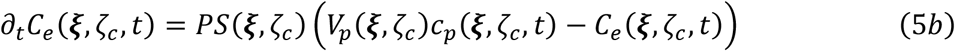

To which we can recover the mass of transport exchange in the voxel

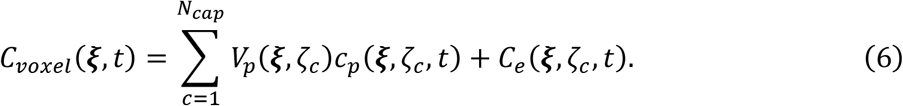

With the total voxel mass determined, the venous tree becomes the next area of focus. We use the capillary concentration *c*_*p*_(***𝝃***, *ζ*_*c*_, *t*) to instantiate the venous terminal concentration *c*_*p*,*vein*_(***𝝃***_***term***_, *t*):

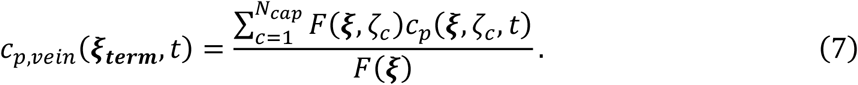

Then, the boundary condition *c*_*p*,*vein*_(***𝝃***_***term***_, *t*) is propagated through the venous tree to obtain *c*_*p*,*vein*_(***𝝃***, *t*) (15,25). Since we have the full geometry of the arterial and venous trees, we have the corresponding arterial volume *V*_*a*_(***𝝃***) and venous volume *V*_*v*_(***𝝃***). The synthetic concentration is then computed:

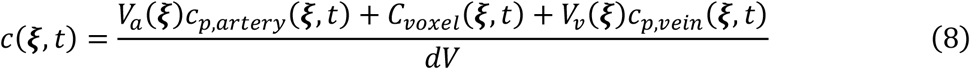

The partial volume fractions *v*_*p*_, *v*_*e*_ can be readily recovered through the relation *V*_*p*_ = *dV* ⋅ *v*_*p*_, *V*_*e*_ = *dV* ⋅ *v*_*e*_. Due to the complementary nature of tissue and tumor ROIs, the concentrations and labels can be combined. For each input cube, there is now a concentration profile *c*(***𝝃***, *t*) with corresponding labels *F*(***𝝃***), *PS*(***𝝃***), *v*_*p*_(***𝝃***) & *v*_*e*_(***𝝃***) sampled at resolution Δ*t* for training a deep neural network (15).

### QTMnet Training

A model was trained to map the synthetic concentration to the underlying parameter maps. For QTMnet, *n* = 250 cubes were used. 150 of those had a mixed tumor and tissue ROI (**Figure 1**), while the remaining 100 cubes were given a uniform ROI. This data was used to train an eleven convolutional layer 3D U-net (See Supplemental figure **S5**). Prior to each training iteration, we apply several augmentations to reduce overfitting. One set of augmentations include x (occurrence rate: 50%), y (occurrence rate: 50%), and z flips (occurrence rate: 50%). To learn the difference between the tissue and background, a masking augmentation was applied to the *tissue* cube ROI as in previous work (15). For a network 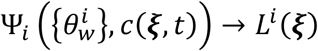 parameterized with weights 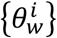 and label *L*^*i*^ ∈ {*F, PS, V*_*p*_, *V*_e_}, the optimal parameters 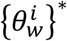 were determined by minimizing the *L*_1_ loss between the network output and label of interest over the 3D volume:

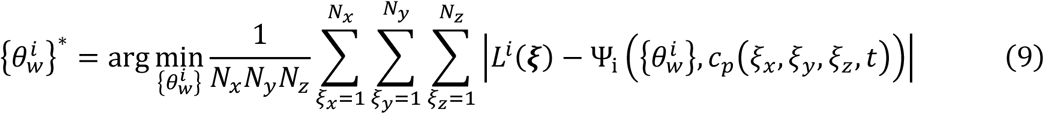

This training problem was optimized with the adaptive moment estimation (ADAM) optimizer (40) with learning rates between 10^−7^ & 10^−6^ and betas (0.9,0.999).

### Two Compartment Exchange Fitting

For comparison, we used the two-compartment exchange (2CXM) model (11):

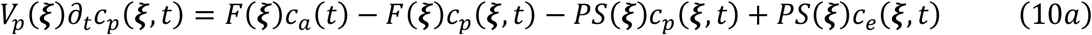

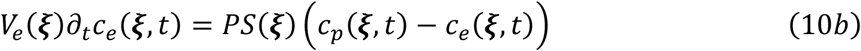

Where *ca*(*t*) is the AIF. We selected the anterior cerebral artery as the AIF location. The 2CXM model can be posed as a linear inversion problem to estimate the desired perfusion parameters (41).

### Data Processing

The DCE-MRI spatiotemporal signal *S*(***𝝃***, *t*) was assumed to be linearly proportional to the Gd concentration:

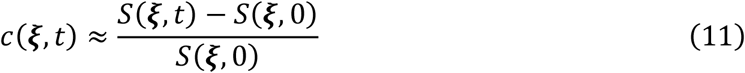

*S*(***𝝃***, 0) is the pre-contrast image. The parameters *F, PS, V*_*p*_, *V*_*e*_ were estimated using QTMnet and 2CXM. Using the tumor ROI, average values are taken for each parameter. A Receiver Operating Characteristic (ROC) curve was constructed for each parameter to determine performance in distinguishing low-from high-grade tumors. We report the area under the curve (AUC) as our performance measure.

### MR Acquisition

30 brain tumor patients were imaged using a *T*_1_ weighted 3D dynamic MRI sequence prior to, during, and after gadolinium injection. The DCE sequence used the following parameters: voxel size = 0.94 × 0.94 × 5*mm*^3^, matrix size = 256x256x*N*_*z*_, *N*_*z*_ ∈ {16,17, …,34} slices, flip angle = 13-25º. TE = 1.21ms, TR = 6.2 ms, temporal resolution = 6.2s, and 24 time points.

### Patient Cohort and ROI definition

This study was approved by the Weill Cornell IRB and all subjects were enrolled via informed consent. 30 patients with suspected glioma were included in this study. Imaging findings and histopathology allowed the tumors to be graded from grade I to grade IV. In this study, 15 tumors were deemed to be ‘low’ grade (grade I or II) and 15 were deemed ‘high’ grade (grade III or IV). For each patient, a tumor ROI was drawn based on the DCE-MRI and *T*_2_-FLAIR imaging findings.

### Results

We provide a representative DCE slice and its corresponding concentration image for the low-grade tumor (**Figure 2A**) We also include *F, PS, V*_*p*_, and *V*_*e*_ maps corresponding to the low-grade lesion (**Figure 3A**). For comparison we also provide a representative high-grade example. This includes the DCE and concentration images (**Figure 2B**), along with the relevant perfusion maps (**Figure 3B**).

**Figure 2.**
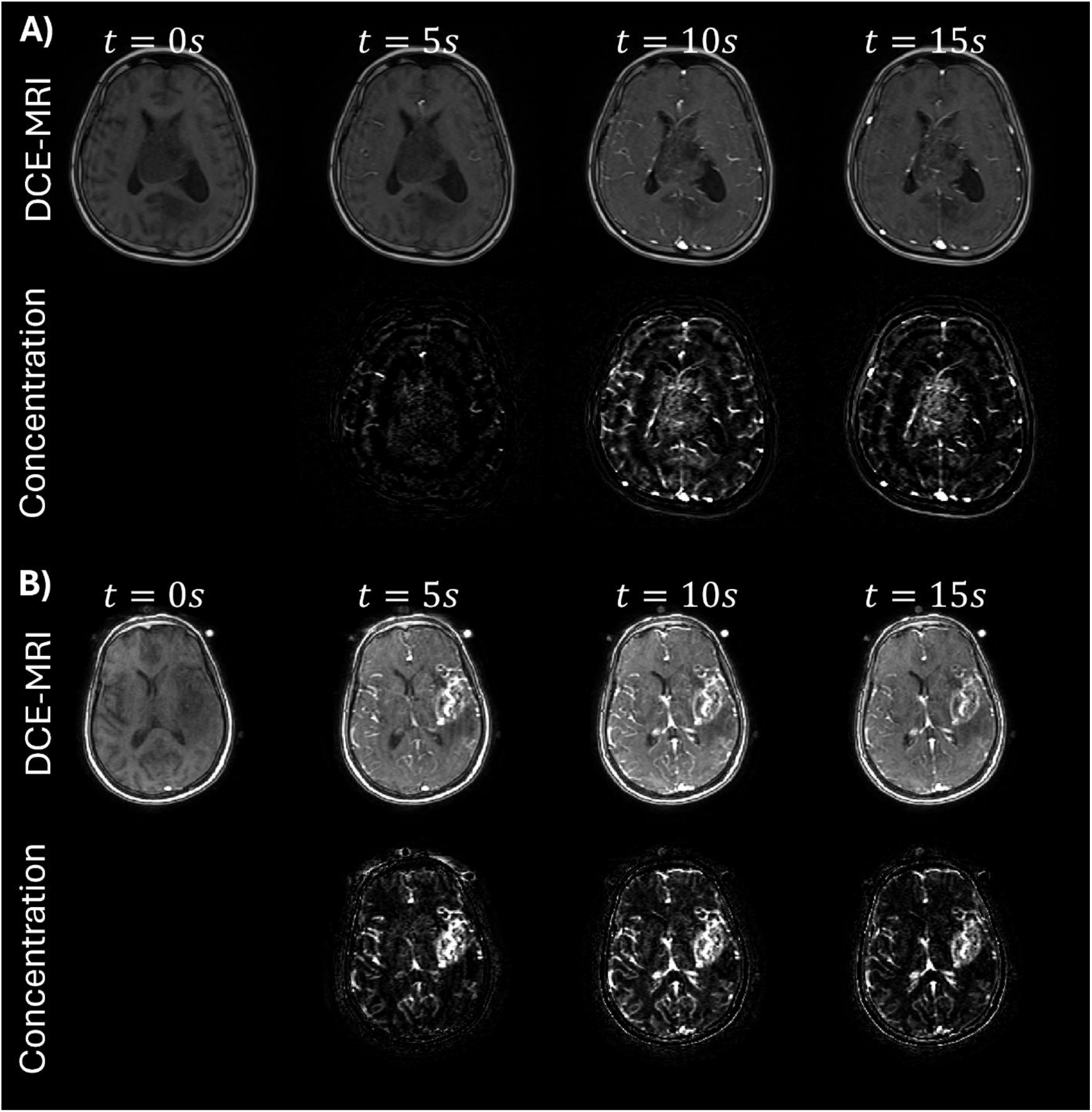
A) Representative DCE-MRI and accompanying concentration image of a low-grade brain tumor. B) Representative DCE-MRI and accompanying concentration image of a high-grade brain tumor.

**Figure 3.**
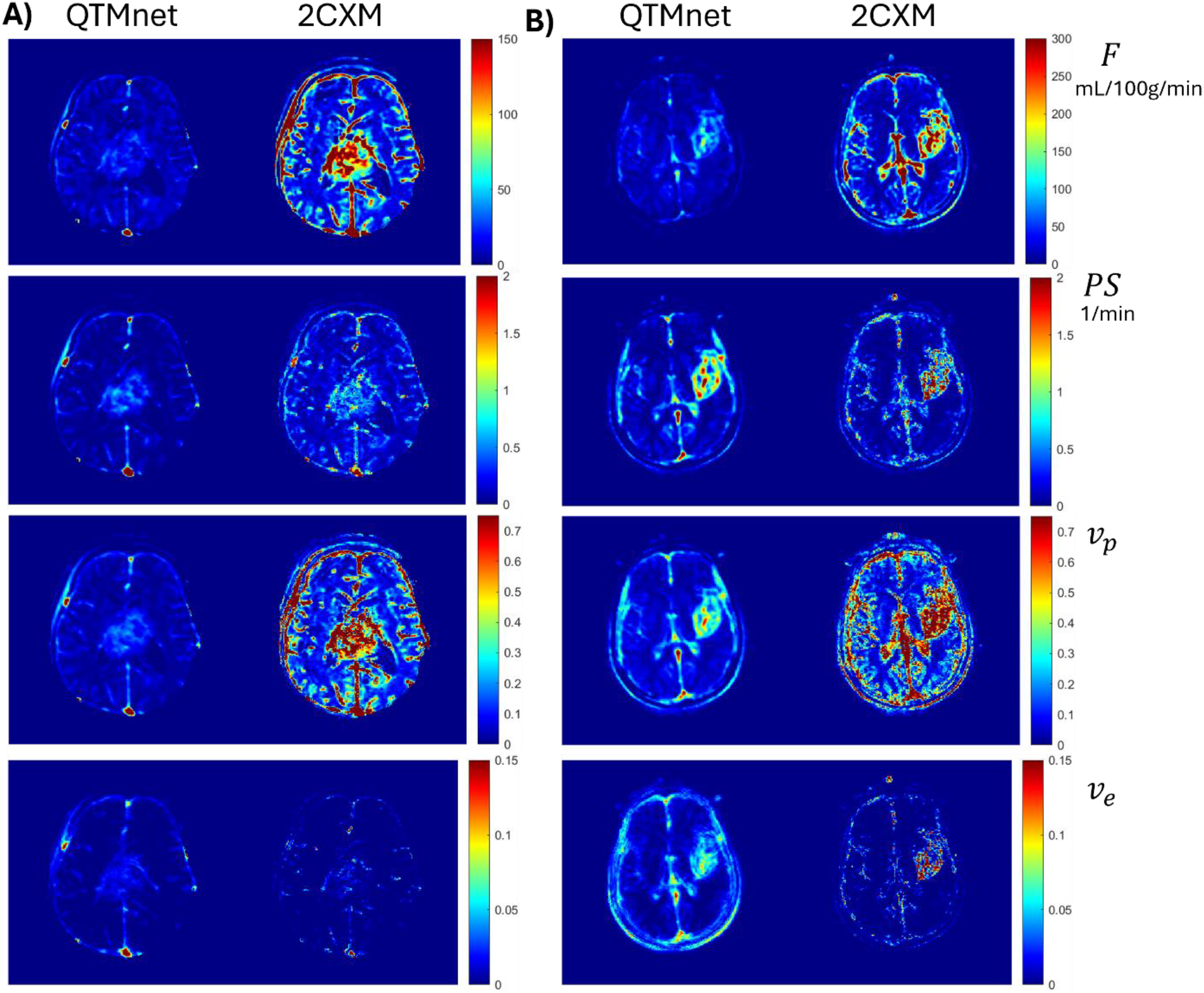
A) Corresponding perfusion map comparisons for the low-grade tumor. B) Corresponding perfusion map comparisons for the high-grade tumor. First (top) tow: F – Flow; Second row: PS – Permeability; Third row: *v*_*p*_ – plasma volume fraction, Fourth (bottom) row: *v*_*e*_ – extravascular extracellular volume fraction.

For the 2CXM inversion, the flow maps produced an AUC of 0.89, PS maps produced an AUC of 0.90, *v*_*p*_ maps produced an AUC of 0.84, and *v*_*e*_ maps produced an AUC of 0.91. For the QTMnet inversion, the flow maps produced an AUC of 0.95, PS maps produced an AUC of 0.97, *v*_*p*_ maps produced an AUC of 0.97, and *v*_*e*_ maps produced an AUC of 0.96. QTMnet consistently outperformed 2CXM in classifying low- and high-grade tumors (**Table 1**). These results can be visualized in the ROC plots (**Figure 4**).

**Table 1.** AUC comparison of two compartment exchange (2CXM), Tissue QTMnet ablation (QTMnet_tissue_), and QTMnet_tumor_.

| *Model* | *F* | *PS* | $v_p$ | $v_e$ |
| --- | --- | --- | --- | --- |
| *2CXM* | 0.89 | 0.90 | 0.84 | 0.91 |
| *QTMnet* | **0.95** | **0.97** | **0.97** | **0.96** |

**Figure 4.**
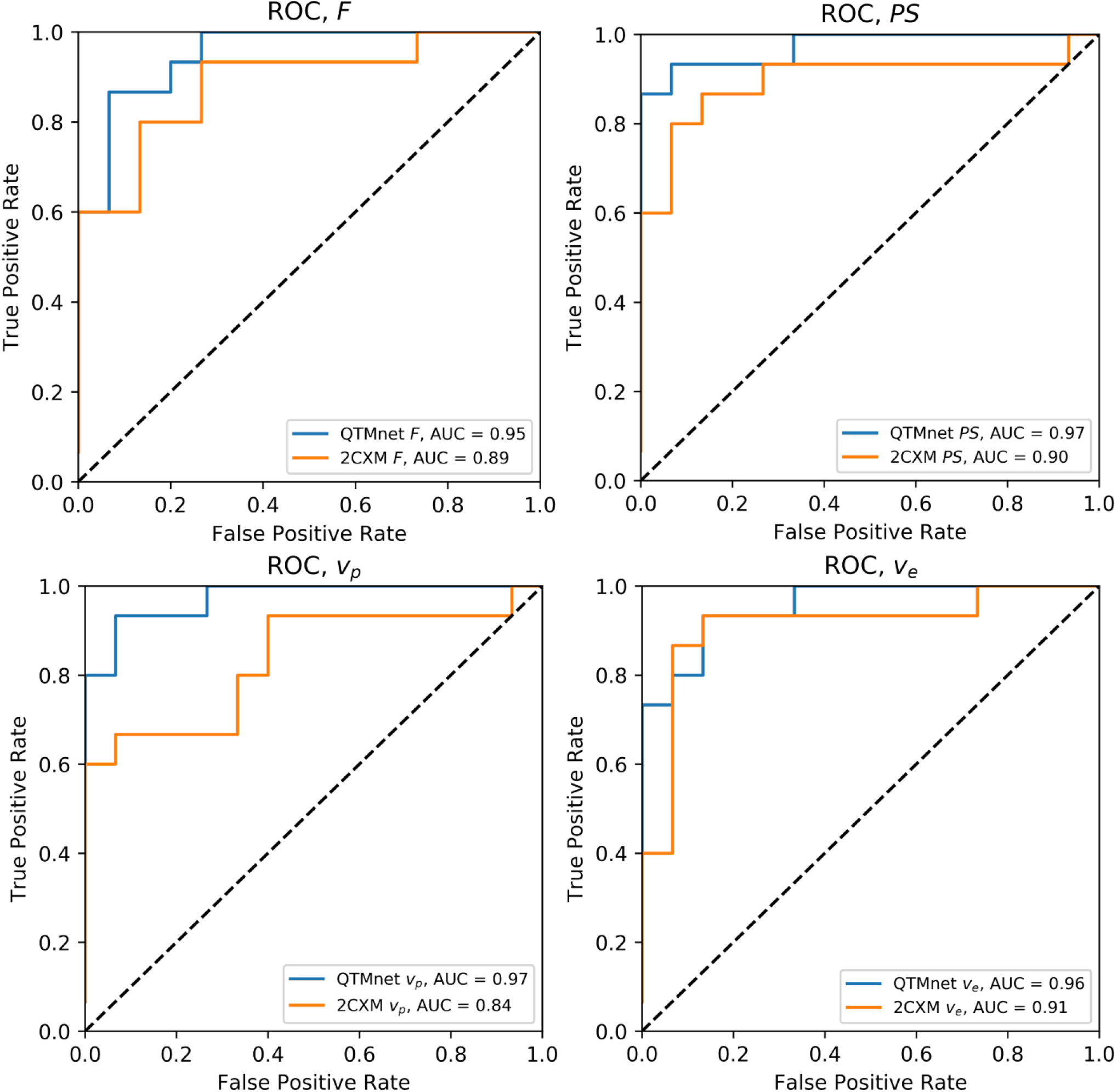
ROC curves. Each compare QTMnet and 2CXM (Top Left) Flow. (Top Right) PS. (Bottom Left) plasma volume fraction. (Bottom Right) Extravascular extracellular volume fraction.

## Discussion

This work shows that QTMnet can improve brain tumor classification performance by including the vascular trees, fluid mechanics simulation, and tumor perfusion characteristics. This was demonstrated by applying our method to 30 brains, half of which were low-grade and the remaining half were high-grade. QTMnet AUC was higher than the standard 2CXM estimation across all parameters.

The QTMnet data generation pipeline ensures a large domain of perfusion states are fed into the model at training time. This can be attributed to the random label generation, CCO tree growth, and fluid mechanics simulation. Instead of relying on a global AIF, we utilize a broad range of local boundary conditions to simulate the voxel concentration. An additional strength of the generation pipeline is that it allows modifications to consider healthy and diseased perfusion states. For instance, healthy brain tissue has low permeability for Gd; however, glioma and glioblastoma are typically highly permeable to gadolinium. This necessitates the implementation of an exchange model for perfusion simulation, which is an extension to previous work (15). The simulation is further modified to incorporate tumor morphology (**Figure 1**) and the corresponding perfusion states. The lack of an AIF at inference time, combined with the inclusion of tissue and tumor perfusion, may be why QTMnet can outperform the 2CXM estimation.

This work consists of several limitations. First, we only have data from one clinical site and aim to include a cohort from an external site. Second, our sample size of 30 patients is relatively small. Larger sample sizes ensure higher confidence in classification performance (42,43). However, collecting more samples takes substantial time and we will address the sample size issue in follow-up work. Additionally, this work focused on a binary classification problem. We intend to extend the study to multiclass classification according to the WHO definitions (44). Data processing assumed a linear relationship between the DCE-MR signal and Gd concentration. Although several methods use signal equations to estimate *R*_1_(*t*) and its corresponding concentration profile (45-47), our clinical protocol did not include this data. For example, signal equation methods either require a pre-contrast *T*_10_ map (47) or specialized pulse sequence implementations (45). It has been shown that in the absence of a *T*_10_ map, the linear relationship between the DCE-MRI outperforms the signal model based non-linear Gd estimation (48). Finally, this study was only focused on brain glioblastoma. QTMnet can be further extended to study cellular binding of tracer in PET (49,50). Additional extensions of hypo-perfused regions can allow us to simulate and characterize stroke (51) and ischemia (52) throughout the body. We can also consider extending the simulation pipeline to include a dual arterial and portal venous input to study liver perfusion (53,54). This can allow for characterizations of hepatocellular carcinoma (55), along with a potential method to non-invasively estimate lung shunt fraction in these patients (56).

## Conclusions

In this study, we assessed the predictive power of perfusion parameters on glioma grade. QTMnet improves classification performance by replacing the globally selected AIF with a domain randomized fluid mechanics-based training data simulation.

## Supporting Information

This supporting information document provides additional information on drawing the boundary condition, approximating tumor morphology with a region growing algorithm, and details on U-Net implementation.

## Adjusting Region Growing Parameters to Model Tumor Morphology

Our implementation of the region growing algorithm generates a seed point within the image field of view. The voxels adjacent in the x, y, and z directions of the FOV are considered for growth. The adjacent voxels are added as mask voxels according to probability *p*_*grow*_. If the voxel is added, it becomes part of a collection of new voxels for region growth, and the seed voxel is discarded as a growth candidate. In any part of the growth algorithm, it is possible that no voxels are added to the tumor ROI and the algorithm returns the grown ROI. However, if this occurs too early, the tumor ROI may be too small for any kind of meaningful representation of what is observed clinically. This can be seen in **Figure S1**. We found that setting a minimum mask size of *N*_*min*_ = 0.005 ⋅ *N*_*FOV*_ ≈ 163 gave the most consistent results for smaller lesions. The minimum FOV requirement affects how the region growing algorithm handles the case where no new neighbors are added. If the algorithm did not add a new neighbor and the number of added points *n*_*mask*_ < *N*_*min*_, then it is re-run until *N*_*min*_ points were added to the tumor ROI. If no new neighbors are added and *n*_*mask*_ > *N*_*min*_, the algorithm returns the tumor ROI. Setting a stopping criterion *N*_*max*_ is also important for the algorithm. If the algorithm is allowed to grow the tumor ROI too much, then the FOV is saturated by the tumor ROI (**Figure S2)**. It is seen that for *N*_*max*_ ≤ 0.35 ⋅ *N*_*FOV*_ the tumor ROI resembles lesion morphology while staying within the cube FOV. The growth probability *p*_*grow*_ has substantial influence over the tumor ROI shape. If it is too small, the ROI loses any meaningful shape. If it is too large, diamond and diagonal patterns emerge, and the ROI does not look physiological. This is why *p*_*grow*_ is selected to be 0.5 (**Figure S3**). Holes are expected to emerge in the region growing algorithm due to the probabilistic growth mechanism. A dilation operation addresses this issue (**Figure S3)**. An additional dilation is used to create a lesion with a necrotic core.

## Drawing Boundary Condition Parameters Prior to Transport Simulation

For boundary condition generation, a function *f*(*t*, {*ρ*_*i*_}) with several parameters *ρ*_*i*_ is selected to model the bolus boundary condition. Each parameter is drawn from a distribution *ρ*_*i*_∼𝒟_*i*_ and fed into *f*(*t*, {*ρ*_*i*_}). All that’s needed is a timepoint or time array and the boundary bolus can be computed for any desired temporal input. Parker developed a model for bolus function in the brain (39):

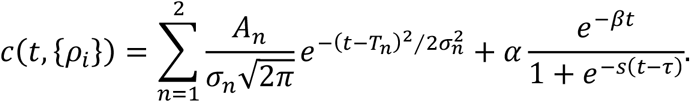

The parameters *ρ*_*i*_ ∈ {*A*_1_, *A*_2_, *T*_1_, *T*_2_, *σ*_1_, *σ*_2_, *α, β, s, τ*} = {*ρ*_1_, *ρ*_2_, *ρ*_3_, *ρ*_4_, *ρ*_5_, *ρ*_6_, *ρ*_7_, *ρ*_8_, *ρ*_9_, *ρ*_10_} are drawn from a normal distribution *ρ*_*i*_∼𝒩(*μ*_*i*_, *σ*_*i*_). Notice that *σ*_*i*_ corresponds to the parameter standard deviation as described in **Table 1**, whereas *σ*_*n*_ corresponds to the ‘spread’ in the bolus first pass (*n* = 1) and second pass (*n* = 2). It is seen that parameters with *n* = 1 refers to the first pass of the bolus, *n* = 2 models the second pass, and the parameters accompanied by *α* models the steady state regime. A comparison between the simulated concentration curves and in-vivo arterial concentrations is provided (**Figure S4**).

## U-Net Architecture

This section provides a description of the model architecture. A summary of the section is found in **Figure S5**. This implementation of the U-Net requires the input concentration to be of shape [1, *t, x, y, z*], so then the 24 input channels correspond to the concentration time points. Since we use 32x32x32 cubes, the numerical dimensions are [1,24,32,32,32]. The first double convolution increases the number of channels to 64 ([1,64,32,32,32]). After the first average pool down sample, the next double convolution doubles the channel number to 128, giving an array shape of [1,128,16,16,16]. A second average pool down-sample brings the data to the information bottleneck (shape: [1,128,8,8,8]). A subsequent convolution doubles the number of channels (shape: [1,256,8,8,8]), and the second convolution reduces the channel number back to 128 (shape: [1,128,8,8,8]). This tensor is spatially up-sampled (shape: [1,128,16,16,16]), concatenated, and fed into a double convolution block to reduce the channel number to [1,64,16,16,16]. Spatial up-sampling ([1,64,32,32,32]) and concatenation ([1,128,32,32,32]) are then repeated once more. A final double convolution reduces to combine the channels into one output channel ([1,1,32,32,32]).

**Figure S1.**
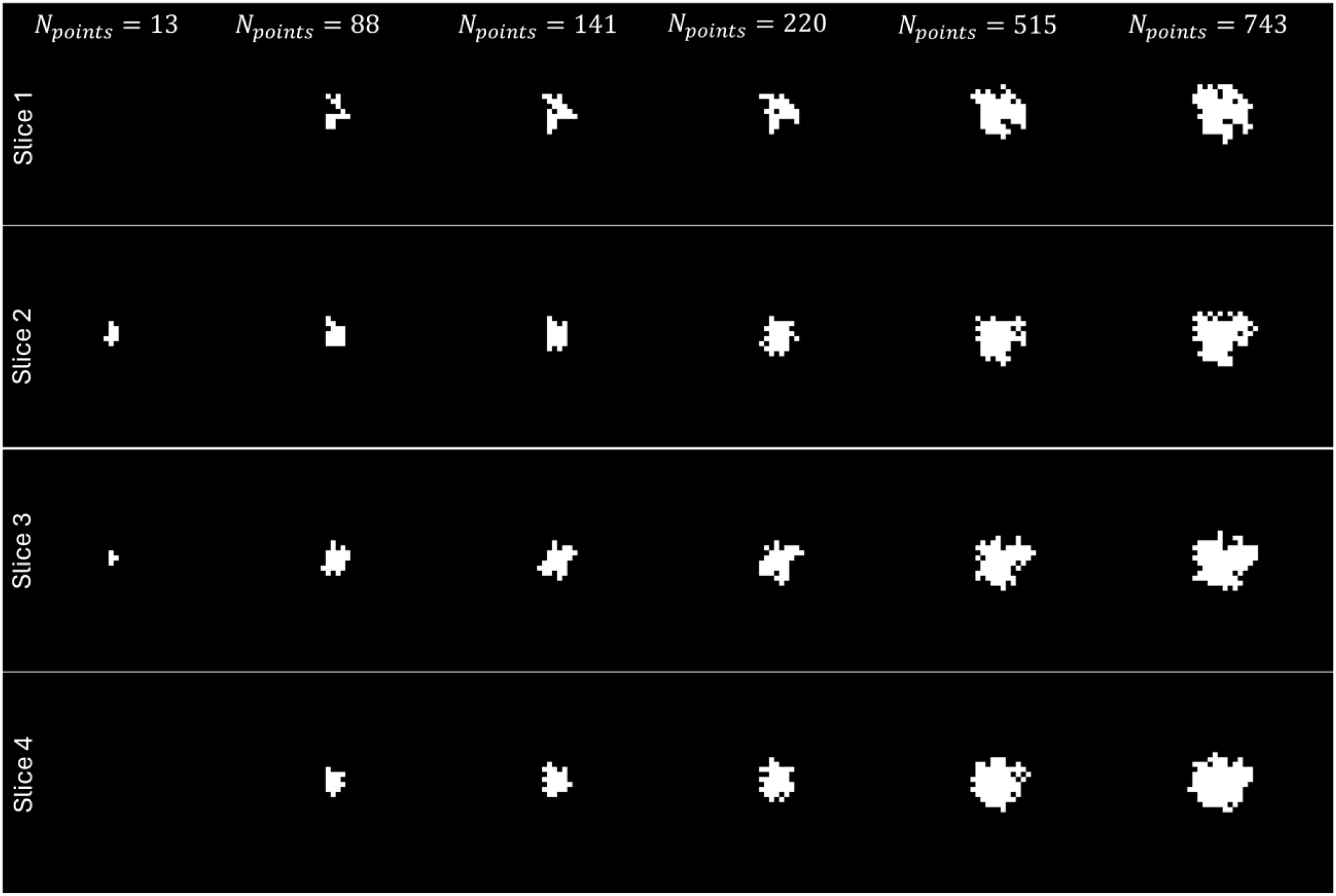
Region grown tumor ROIs with increasing ROI size.

**Figure S2.**
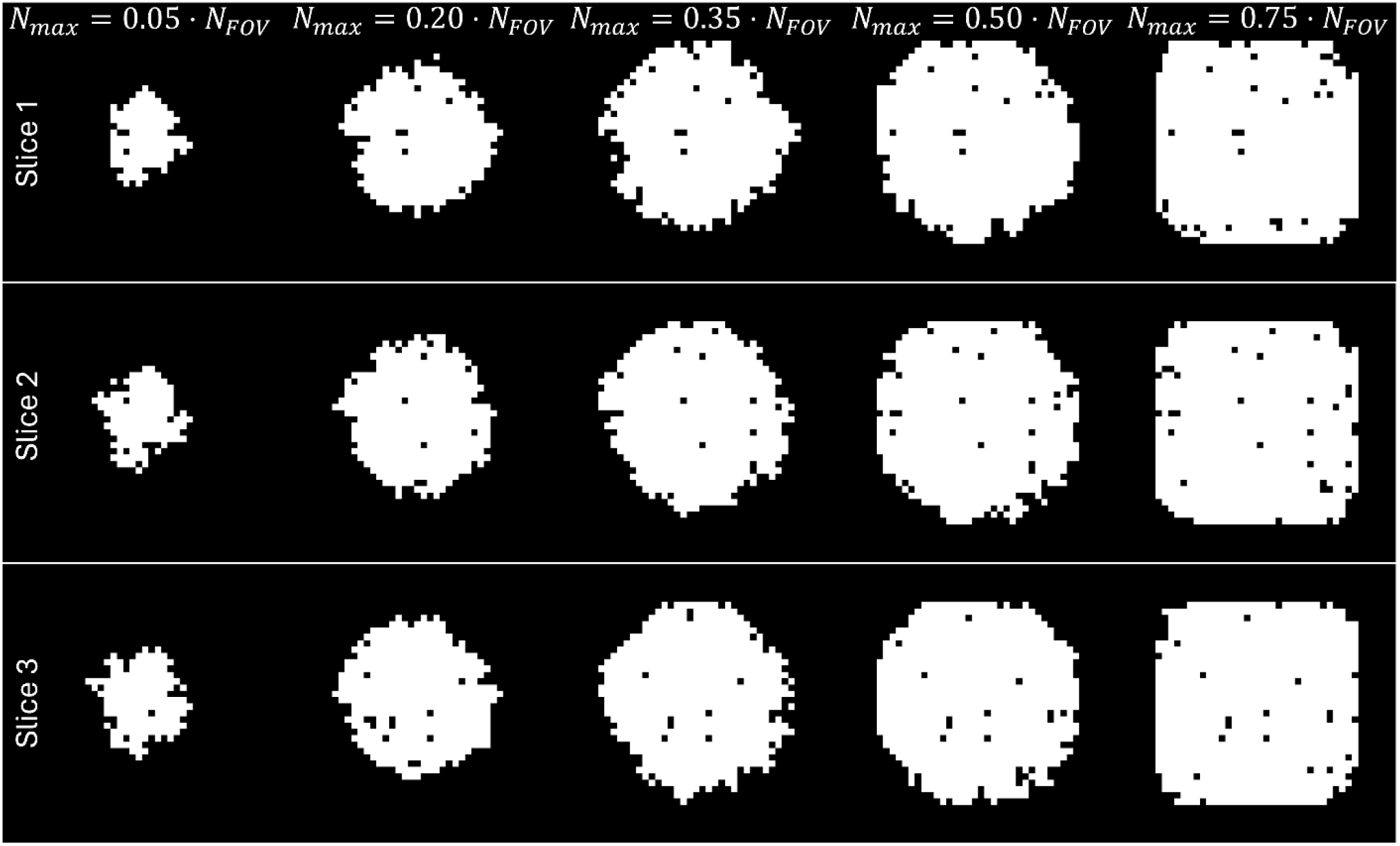
Region Growing for progressively larger stopping criteria.

**Figure S3.**
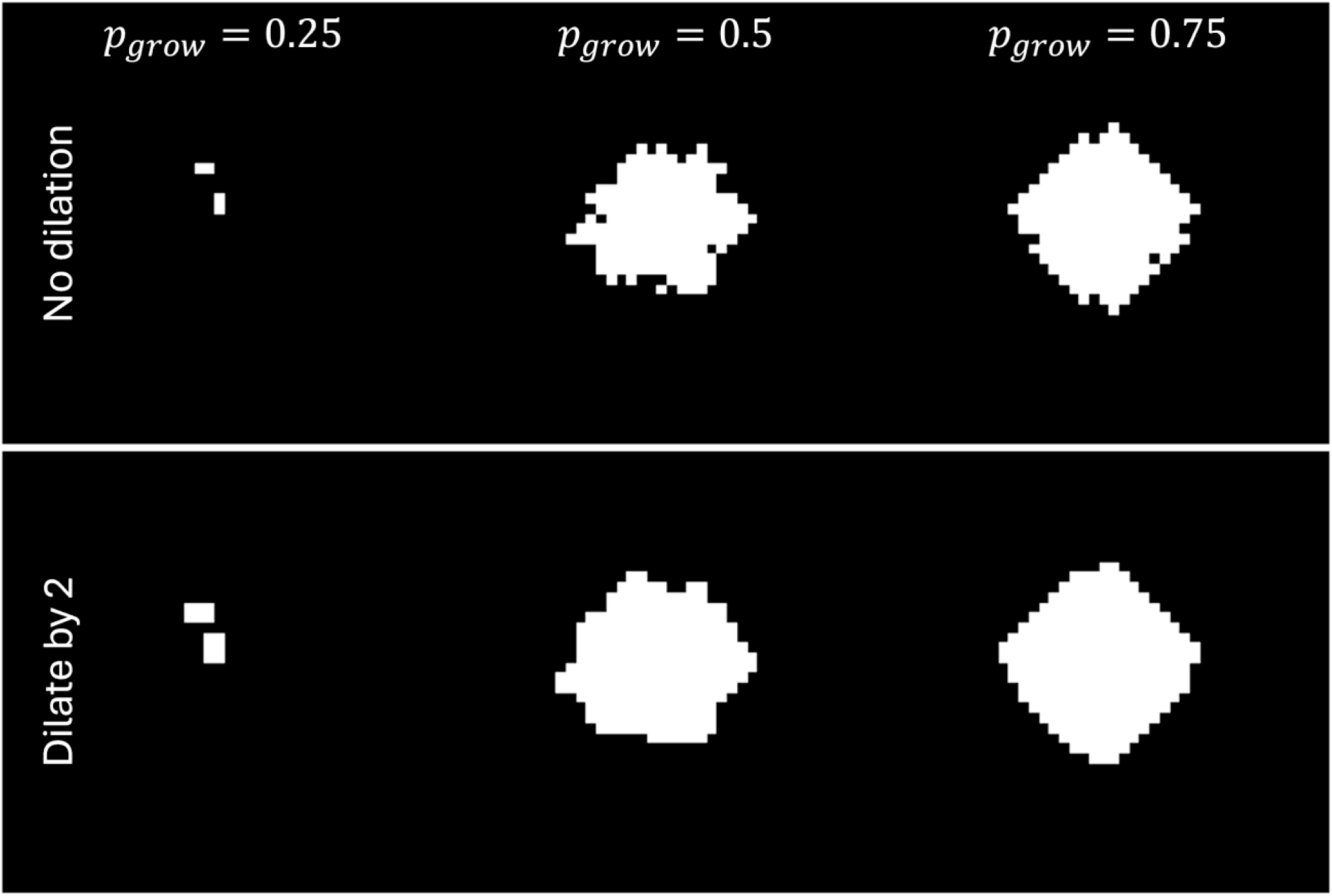
Tumor ROI for different growth probabilities. Dilation is used to remove holes from the mask.

**Table 1.** Parker model parameters used to generate the boundary condition for the tracer simulation. Each parameter is drawn from the Gaussian distribution *param*∼𝒩(*mean, SD*^2^).

| *Parameter* | $A_1$ | $A_2$ | $T_1$ | $T_2$ | $\sigma_1$ | $\sigma_2$ | $\alpha$ | $\beta$ | $s$ | $\tau$ |
| --- | --- | --- | --- | --- | --- | --- | --- | --- | --- | --- |
| *Mean* | 46.1 | 23.8 | 10.22 | 21.86 | 3.38 | 7.92 | 1.27 | $5.86 \cdot 10^{-3}$ | 0.636 | 29.0 |
| *SD* | 21.2 | 11.8 | 0.044 | 1.66 | 0.066 | 1.27 | 1.84 | $3.24 \cdot 10^{-3}$ | 0.282 | 0.91 |
| *Unit* | *mmol*<br>*· sec* | *mmol*<br>*· sec* | *sec* | *sec* | *mmol*<br>*· sec* | *mmol*<br>*· sec* | *mmol* | *sec*<sup>-1</sup> | *sec*<sup>-1</sup> | *sec* |

**Figure S4.**
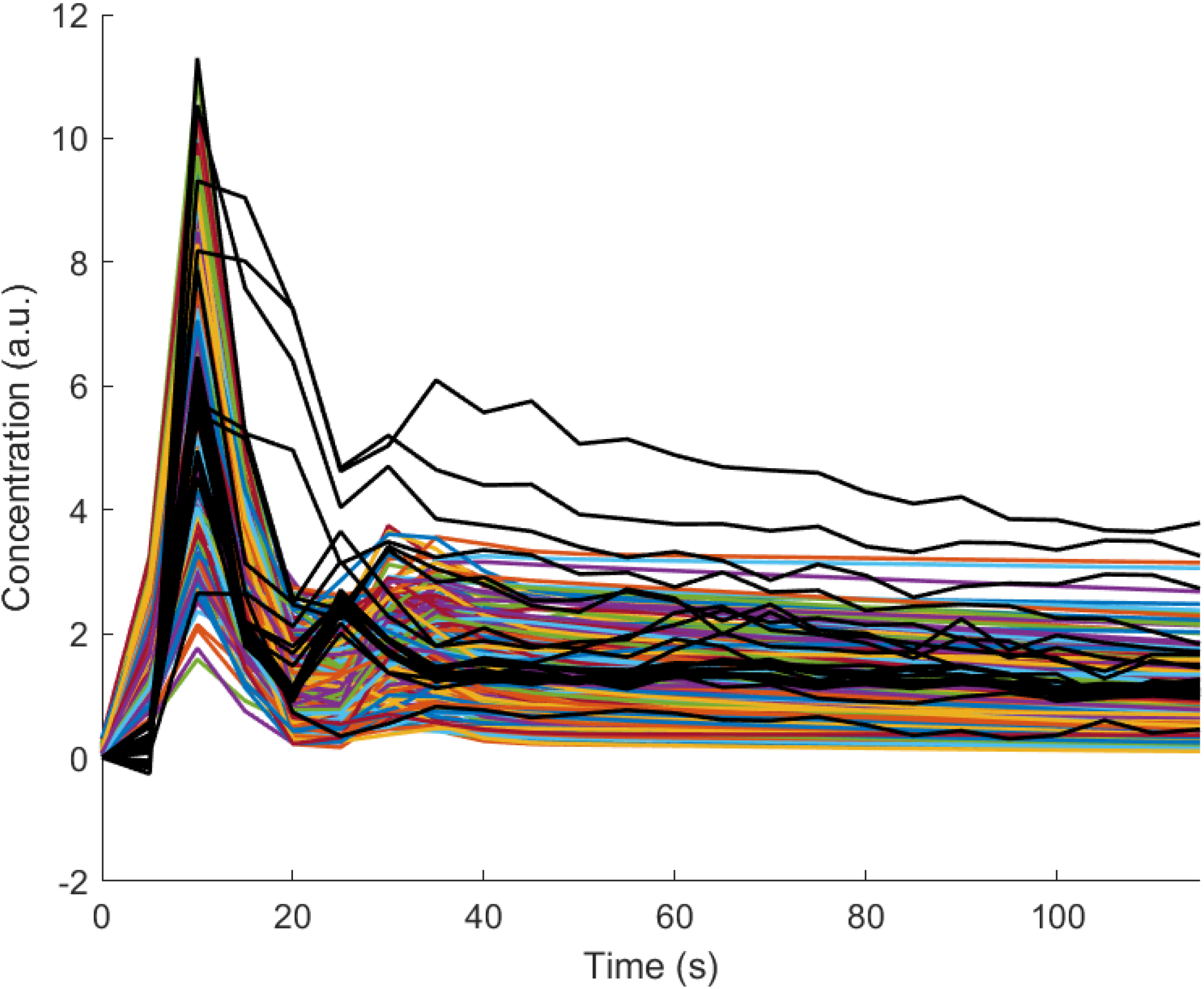
A comparison between the simulated Parker model curves (multi-colored) and anterior cerebral artery concentrations.

**Figure S5.**
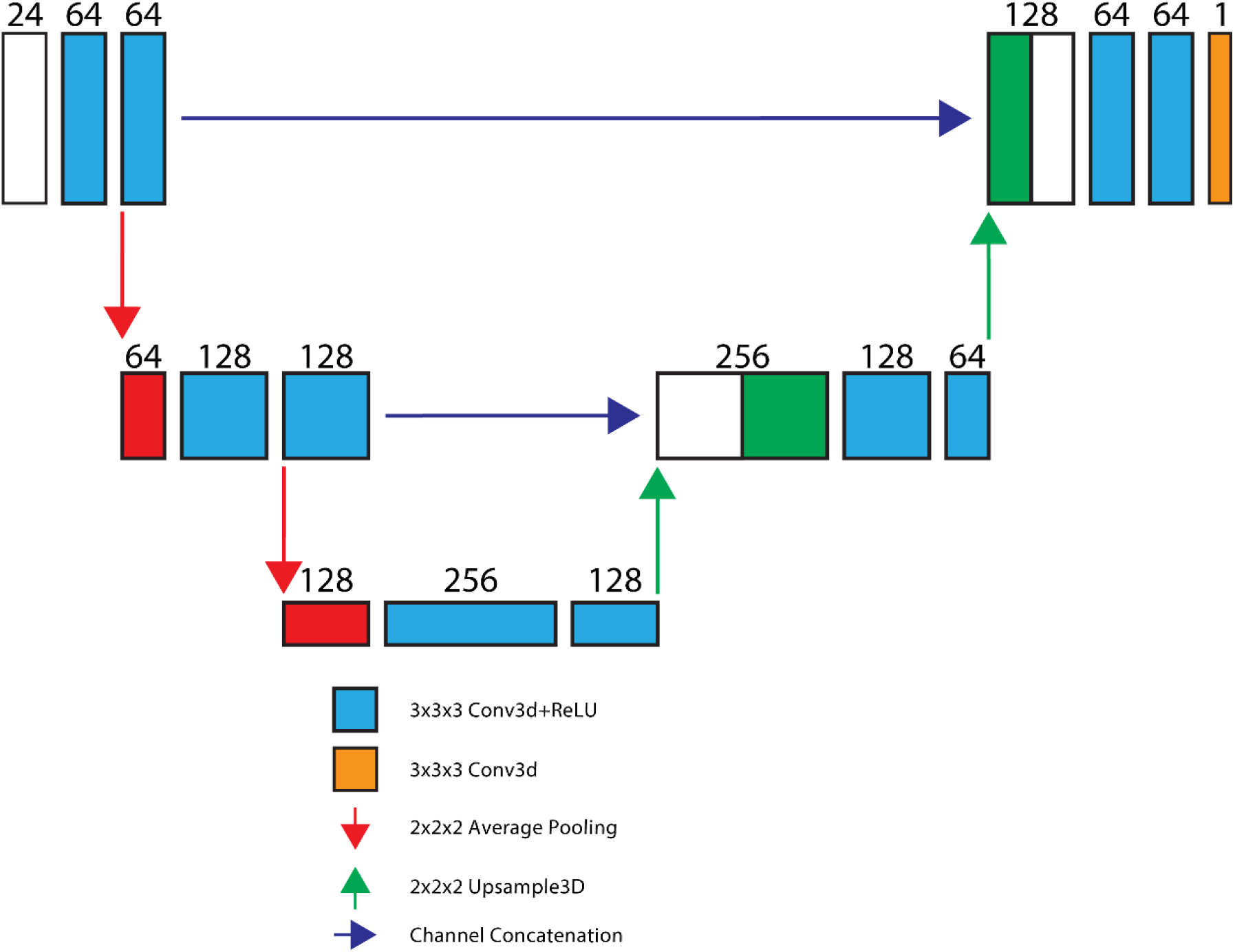
“U-Net” model with eleven convolutional layers. Input dimensions are 32x32x32 voxels with 24 input channels. The input channels correspond to the number of time-points in the DCE-MRI data.

## Data Availability

All data produced in the present work are contained in the manuscript

## References

1. Hanif F, Muzaffar K, Perveen K, Malhi SM, Simjee Sh U. Glioblastoma Multiforme: A Review of its Epidemiology and Pathogenesis through Clinical Presentation and Treatment. Asian Pac J Cancer Prev 2017;18(1):3–9.

2. Silantyev AS, Falzone L, Libra M, Gurina OI, Kardashova KS, Nikolouzakis TK, Nosyrev AE, Sutton CW, Mitsias PD, Tsatsakis A. Current and Future Trends on Diagnosis and Prognosis of Glioblastoma: From Molecular Biology to Proteomics. Cells 2019;8(8):863.

3. Hishii M, Matsumoto T, Arai H. Diagnosis and Treatment of Early-Stage Glioblastoma. Asian J Neurosurg 2019;14(2):589–592.

4. Kawauchi D, Ohno M, Miyakita Y, Takahashi M, Yanagisawa S, Omura T, Yoshida A, Kubo Y, Igaki H, Ichimura K, Narita Y. Early Diagnosis and Surgical Intervention Within 3 Weeks From Symptom Onset Are Associated With Prolonged Survival of Patients With Glioblastoma. Neurosurgery 2022;91(5):741–748.

5. Arevalo-Perez J, Peck KK, Young RJ, Holodny AI, Karimi S, Lyo JK. Dynamic Contrast-Enhanced Perfusion MRI and Diffusion-Weighted Imaging in Grading of Gliomas. J Neuroimaging 2015;25(5):792–798.

6. Desjardins A, Barboriak DP, Herndon JE, Reardon DA, Quinn JA, Rich JN, Sathornsumetee S, Gururangan S, Friedman HS, Vredenburgh JJ. Dynamic contrast-enhanced magnetic resonance imaging (DCE-MRI) evaluation in glioblastoma (GBM) patients treated with bevacizumab (BEV) and irinotecan (CPT-11). Journal of Clinical Oncology 2007;25(18_suppl):2029–2029.

7. Vankan B, Var V, Rommers N, Psychogios MN, Todea RA. Treatment response assessment in glioblastoma with GRASP DCE-MRI. European Journal of Radiology 2025;190:112234.

8. Zerweck L, Hauser T-K, Klose U, Han T, Nägele T, Shen M, Gohla G, Estler A, Xie C, Hu H, Yang S, Cao Z, Erb G, Ernemann U, Richter V. Glioma Type Prediction with Dynamic Contrast-Enhanced MR Imaging and Diffusion Kurtosis Imaging—A Standardized Multicenter Study. Cancers 2024;16(15):2644.

9. Okuchi S, Rojas-Garcia A, Ulyte A, Lopez I, Ušinskienė J, Lewis M, Hassanein SM, Sanverdi E, Golay X, Thust S, Panovska-Griffiths J, Bisdas S. Diagnostic accuracy of dynamic contrast-enhanced perfusion MRI in stratifying gliomas: A systematic review and meta-analysis. Cancer Med 2019;8(12):5564–5573.

10. Tofts PS, Kermode AG. Measurement of the blood-brain barrier permeability and leakage space using dynamic MR imaging. 1. Fundamental concepts. Magn Reson Med 1991;17(2):357–367.

11. Sourbron SP, Buckley DL. On the scope and interpretation of the Tofts models for DCE-MRI. Magnetic Resonance in Medicine 2011;66(3):735–745.

12. Sourbron SP, Buckley DL. Classic models for dynamic contrast-enhanced MRI. NMR in Biomedicine 2013;26(8):1004–1027.

13. Zhou L, Zhang Q, Spincemaille P, Nguyen TD, Morgan J, Dai W, Li Y, Gupta A, Prince MR, Wang Y. Quantitative transport mapping (QTM) of the kidney with an approximate microvascular network. Magnetic Resonance in Medicine 2021;85(4):2247–2262.

14. Calamante F. Bolus dispersion issues related to the quantification of perfusion MRI data. J Magn Reson Imaging 2005;22(6):718–722.

15. Romano D, Zhang Q, Roberts AG, Weppner B, Shou J, Kovanlikaya I, Şişman M, Hu R, Nguyen TD, Dimov AV, Prince MR, Spincemaille P, Wang Y. Validation of quantitative transport mapping (QTM) with an ex vivo perfused liver model. Magnetic Resonance in Medicine 2025;94(4):1779–1792.

16. Calamante F. Arterial input function in perfusion MRI: a comprehensive review. Prog Nucl Magn Reson Spectrosc 2013;74:1–32.

17. Keil VC, Mädler B, Gieseke J, Fimmers R, Hattingen E, Schild HH, Hadizadeh DR. Effects of arterial input function selection on kinetic parameters in brain dynamic contrast-enhanced MRI. Magn Reson Imaging 2017;40:83–90.

18. Liu P, Lee YZ, Aylward SR, Niethammer M. Perfusion Imaging: An Advection Diffusion Approach. IEEE Transactions on Medical Imaging 2021;40(12):3424–3435.

19. Sourbron S. A Tracer-Kinetic Field Theory for Medical Imaging. IEEE Transactions on Medical Imaging 2014;33(4):935–946.

20. Zhou L, Zhang Q, Spincemaille P, Nguyen T, Morgan J, Dai W, Gupta A, Prince MR, Wang Y. Perfusion Quantification Validation on a Numerical Vascular Network of the Kidney: Traditional Kety’s Method vs Quantitative Transport Mapping. 2020. ISMRM.

21. Liu P, Lee YZ, Aylward SR, Niethammer M. PIANO: Perfusion Imaging via Advection-Diffusion. Medical Image Computing and Computer Assisted Intervention – MICCAI 2020; 2020; Cham. Springer International Publishing. p 688–698. (Medical Image Computing and Computer Assisted Intervention – MICCAI 2020).

22. Zhang Q, Luo X, Zhou L, Nguyen TD, Prince MR, Spincemaille P, Wang Y. Fluid Mechanics Approach to Perfusion Quantification: Vasculature Computational Fluid Dynamics Simulation, Quantitative Transport Mapping (QTM) Analysis of Dynamics Contrast Enhanced MRI, and Application in Nonalcoholic Fatty Liver Disease Classification. IEEE Transactions on Biomedical Engineering 2023;70(3):980–990.

23. Zhang Q, Romano D, Weppner B, Nguyen T, Spincemaille P, Wang Y. Fluid mechanics based quantitative transport mapping network for predicting gene expression of nasopharyngeal carcinoma (NPC) patients. ISMRM Annual Meeting: ISMRM; 2024.

24. Zhang Q, Spincemaille P, Drotman M, Chen C, Eskreis-Winkler S, Huang W, Zhou L, Morgan J, Nguyen TD, Prince MR, Wang Y. Quantitative transport mapping (QTM) for differentiating benign and malignant breast lesion: Comparison with traditional kinetics modeling and semi-quantitative enhancement curve characteristics. Magn Reson Imaging 2022;86:86–93.

25. Zhang Q. Estimating Perfusion and Vascular Properties from Medical Images: Quantitative Transport Mapping (QTM) [Ph.D.]. United States -- New York: Cornell University; 2023. 104 p.

26. Zhang Q, Romano D, Nguyen T, Spincemaille P, Wang Y. Estimating perfusion and permeability using neural network with training data generated from vessel construction and transport simulation. ISMRM Annual Meeting: ISMRM; 2023.

27. D. Romano QZ, M. Sisman, A. Roberts, R. Hu, B. Weppner, T. Nguyen, P. Spincemaille, M. Prince, Y. Wang. Validating Quantitative Transport Mapping (QTM) on a Perfused MRI Liver Phantom. In: Wiley, editor 2024. Magnetic Resonance in Medicine.

28. Sankaranarayanan S, Balaji Y, Jain A, Lim SN, Chellappa R. Learning from Synthetic Data: Addressing Domain Shift for Semantic Segmentation. 2018 IEEE/CVF Conference on Computer Vision and Pattern Recognition (CVPR): IEEE Computer Society; 2018. p 3752–3761.

29. Tremblay J, Prakash A, Acuna D, Brophy M, Jampani V, Anil C, To T, Cameracci E, Boochoon S, Birchfield S. Training deep networks with synthetic data: Bridging the reality gap by domain randomization. 2018. p 969–977.

30. Billot B, Greve DN, Puonti O, Thielscher A, Van Leemput K, Fischl B, Dalca AV, Iglesias JE. SynthSeg: Segmentation of brain MRI scans of any contrast and resolution without retraining. Medical Image Analysis 2023;86:102789.

31. Hoffmann M, Billot B, Greve DN, Iglesias JE, Fischl B, Dalca AV. SynthMorph: Learning Contrast-Invariant Registration Without Acquired Images. IEEE Transactions on Medical Imaging 2022;41(3):543–558.

32. Hu R, Zhang Q, Romano D, Wang Y. Quantitative transport mapping (QTM) of the brain with simulated microvasculature model. ISMRM Annual Meeting: ISMRM; 2024.

33. Karch R, Neumann F, Neumann M, Schreiner W. A three-dimensional model for arterial tree representation, generated by constrained constructive optimization. Comput Biol Med 1999;29(1):19–38.

34. Schreiner W. Computer generation of complex arterial tree models. Journal of Biomedical Engineering 1993;15(2):148–150.

35. Xing CY, Tarumi T, Liu J, Zhang Y, Turner M, Riley J, Tinajero CD, Yuan LJ, Zhang R. Distribution of cardiac output to the brain across the adult lifespan. J Cereb Blood Flow Metab 2017;37(8):2848–2856.

36. Cosgrove KP, Mazure CM, Staley JK. Evolving knowledge of sex differences in brain structure, function, and chemistry. Biol Psychiatry 2007;62(8):847–855.

37. Lu SS, Cao YZ, Su CQ, Xu XQ, Zhao LB, Jia ZY, Liu QH, Hsu YC, Liu S, Shi HB, Wu FY. Hyperperfusion on Arterial Spin Labeling MRI Predicts the 90-Day Functional Outcome After Mechanical Thrombectomy in Ischemic Stroke. J Magn Reson Imaging 2021;53(6):1815–1822.

38. Zhao M, Guo LL, Huang N, Wu Q, Zhou L, Zhao H, Zhang J, Fu K. Quantitative analysis of permeability for glioma grading using dynamic contrast-enhanced magnetic resonance imaging. Oncol Lett 2017;14(5):5418–5426.

39. Parker GJ, Roberts C, Macdonald A, Buonaccorsi GA, Cheung S, Buckley DL, Jackson A, Watson Y, Davies K, Jayson GC. Experimentally-derived functional form for a population-averaged high-temporal-resolution arterial input function for dynamic contrast-enhanced MRI. Magn Reson Med 2006;56(5):993–1000.

40. Kingma DP, Ba J. Adam: A method for stochastic optimization. arXiv [csLG] 2014.

41. Flouri D, Lesnic D, Sourbron SP. Fitting the two-compartment model in DCE-MRI by linear inversion. Magnetic Resonance in Medicine 2016;76(3):998–1006.

42. Figueroa RL, Zeng-Treitler Q, Kandula S, Ngo LH. Predicting sample size required for classification performance. BMC Medical Informatics and Decision Making 2012;12(1):8.

43. Xu J. On the bias in the AUC variance estimate. Pattern Recognition Letters 2024;178:62–68.

44. Kurokawa R, Kurokawa M, Baba A, Ota Y, Pinarbasi E, Camelo-Piragua S, Capizzano AA, Liao E, Srinivasan A, Moritani T. Major Changes in 2021 World Health Organization Classification of Central Nervous System Tumors. RadioGraphics 2022;42(5):1474–1493.

45. Bharadwaj Das A, Tranos JA, Zhang J, Zaim Wadghiri Y, Kim SG. Estimation of Contrast Agent Concentration in DCE-MRI Using 2 Flip Angles. Investigative Radiology 2022;57(5):343–351.

46. Shalom ES, Kim H, van der Heijden RA, Ahmed Z, Patel R, Hormuth II DA, DiCarlo JC, Yankeelov TE, Sisco NJ, Dortch RD, Stokes AM, Inglese M, Grech-Sollars M, Toschi N, Sahoo P, Singh A, Verma SK, Rathore DK, Kazerouni AS, Partridge SC, LoCastro E, Paudyal R, Wolansky IA, Shukla-Dave A, Schouten P, Gurney-Champion OJ, Jirík R, Macícek O, Bartoš M, Vitouš J, Das AB, Kim SG, Bokacheva L, Mikheev A, Rusinek H, Berks M, Hubbard Cristinacce PL, Little RA, Cheung S, O’Connor JPB, Parker GJM, Moloney B, LaViolette PS, Bobholz S, Duenweg S, Virostko J, Laue HO, Sung K, Nabavizadeh A, Saligheh Rad H, Hu LS, Sourbron S, Bell LC, Fathi Kazerooni A. The ISMRM Open Science Initiative for Perfusion Imaging (OSIPI): Results from the OSIPI–Dynamic Contrast-Enhanced challenge. Magnetic Resonance in Medicine 2024;91(5):1803–1821.

47. Hittmair K, Gomiscek G, Langenberger K, Recht M, Imhof H, Kramer J. Method for the quantitative assessment of contrast agent uptake in dynamic contrast-enhanced MRI. Magnetic Resonance in Medicine 1994;31(5):567–571.

48. Wake N, Chandarana H, Rusinek H, Fujimoto K, Moy L, Sodickson DK, Kim SG. Accuracy and precision of quantitative DCE-MRI parameters: How should one estimate contrast concentration? Magn Reson Imaging 2018;52:16–23.

49. Lammertsma AA, Bench CJ, Hume SP, Osman S, Gunn K, Brooks DJ, Frackowiak RS. Comparison of methods for analysis of clinical [11C]raclopride studies. J Cereb Blood Flow Metab 1996;16(1):42–52.

50. Lammertsma AA, Hume SP. Simplified reference tissue model for PET receptor studies. Neuroimage 1996;4(3 Pt 1):153–158.

51. Carpenter TK, Armitage PA, Bastin ME, Wardlaw JM. DSC perfusion MRI-Quantification and reduction of systematic errors arising in areas of reduced cerebral blood flow. Magn Reson Med 2006;55(6):1342–1349.

52. Sliwicka O, Sechopoulos I, Baggiano A, Pontone G, Nijveldt R, Habets J. Dynamic myocardial CT perfusion imaging-state of the art. Eur Radiol 2023;33(8):5509–5525.

53. Aronhime S, Calcagno C, Jajamovich GH, Dyvorne HA, Robson P, Dieterich D, Fiel MI, Martel-Laferriere V, Chatterji M, Rusinek H, Taouli B. DCE-MRI of the liver: effect of linear and nonlinear conversions on hepatic perfusion quantification and reproducibility. J Magn Reson Imaging 2014;40(1):90–98.

54. Jafari R, Chhabra S, Prince MR, Wang Y, Spincemaille P. Vastly accelerated linear least-squares fitting with numerical optimization for dual-input delay-compensated quantitative liver perfusion mapping. Magn Reson Med 2018;79(4):2415–2421.

55. Chen BB, Shih TT. DCE-MRI in hepatocellular carcinoma-clinical and therapeutic image biomarker. World J Gastroenterol 2014;20(12):3125–3134.

56. Mehadji B, Marx T, Carter A, Goldman RE, Vu CT, Roncali E. Contrast-enhanced CT as a non-invasive alternative for lung shunt fraction estimation in hepatic transarterial radioembolization. Radiol Adv 2025;2(4):umaf025.

